# The *LRRK2* R1441G+M1646T haplotype is associated with slower motor symptom progression in Parkinson’s disease

**DOI:** 10.64898/2026.08.26.26361428

**Authors:** Jackson G. Schumacher, Xinyuan Zhang, Jian Wang, Xiqun Chen

**Author notes:** Correspondence to: Xiqun Chen, Department of Neurology, Massachusetts General Hospital, and Harvard Medical School, Boston, MA 02129, USA.

## Abstract

**Background:** Mutations in leucine-rich repeat kinase 2 (*LRRK2*) are the most common genetic risk factor for Parkinson’s disease (PD). G2019S, the most common pathogenic variant, has been linked to milder motor symptoms, but the effects of other LRRK2 variants on disease trajectory remain incompletely characterized. R1441G, the second most common pathogenic variant, co-occurs with the PD risk variant M1646T on a shared haplotype. Whether this haplotype confers a distinct rate of motor progression has not been established.

**Methods:** We analyzed up to 12 years of longitudinal data from 603 participants in the Parkinson’s Progression Markers Initiative (PPMI) with PD and available whole-genome sequencing data: 394 sporadic PD, 169 G2019S carriers, 20 R1441G+M1646T carriers, and 20 M1646T carriers. Motor symptom progression (MDS-UPDRS III) was assessed using linear mixed-effects models with genotype-by-time interactions, adjusted for age at onset, disease duration at baseline, sex, race, baseline score, and levodopa equivalent daily dose.

**Results:** R1441G+M1646T carriers exhibited 76% slower progression in OFF-state MDS-UPDRS III than sporadic PD (0.50 vs. 2.04 points/year; β=−1.54 [95% CI: −2.48, −0.60]; p=0.001). G2019S carriers exhibited 26% slower progression (1.52 points/year; β=−0.52 [−0.99, −0.06]; p=0.03). M1646T carriers did not differ from sporadic PD (p=0.60). Slower progression in R1441G+M1646T carriers was characterized by attenuated bradykinesia (64% slower; p=0.008), axial decline (76% slower; p=0.002), and a lack of orofacial symptom progression (p<0.001). R1441G+M1646T carriers also exhibited 55% slower self-reported motor decline (MDS-UPDRS II; p=0.04)

**Conclusions:** R1441G+M1646T carriers exhibit substantially slower motor progression than sporadic PD while M1646T carriers do not.

## Introduction

Mutations in leucine-rich repeat kinase 2 (*LRRK2*) are the most common genetic risk factor for Parkinson’s disease (PD).^1^ Pathogenic variants span distinct functional domains of the LRRK2 protein influencing GTPase and kinase activity (Figure 1a).^1,2^ G2019S, the most common pathogenic *LRRK2* variant, has been linked to milder motor symptoms.^2^ R1441G, the second most common pathogenic variant, co-occurs with the PD risk variant M1646T on a shared haplotype, yet whether this haplotype confers a distinct clinical trajectory has not been investigated.^1–3^ While M1646T can occur independently, R1441G has not been reported independently of M1646T.^1,4,5^ Using longitudinal data from the Parkinson’s Progression Markers Initiative (PPMI), we compared motor and non-motor symptom progression across G2019S carriers, R1441G+M1646T haplotype carriers, and M1646T carriers relative to sporadic PD.

**Figure 1.**
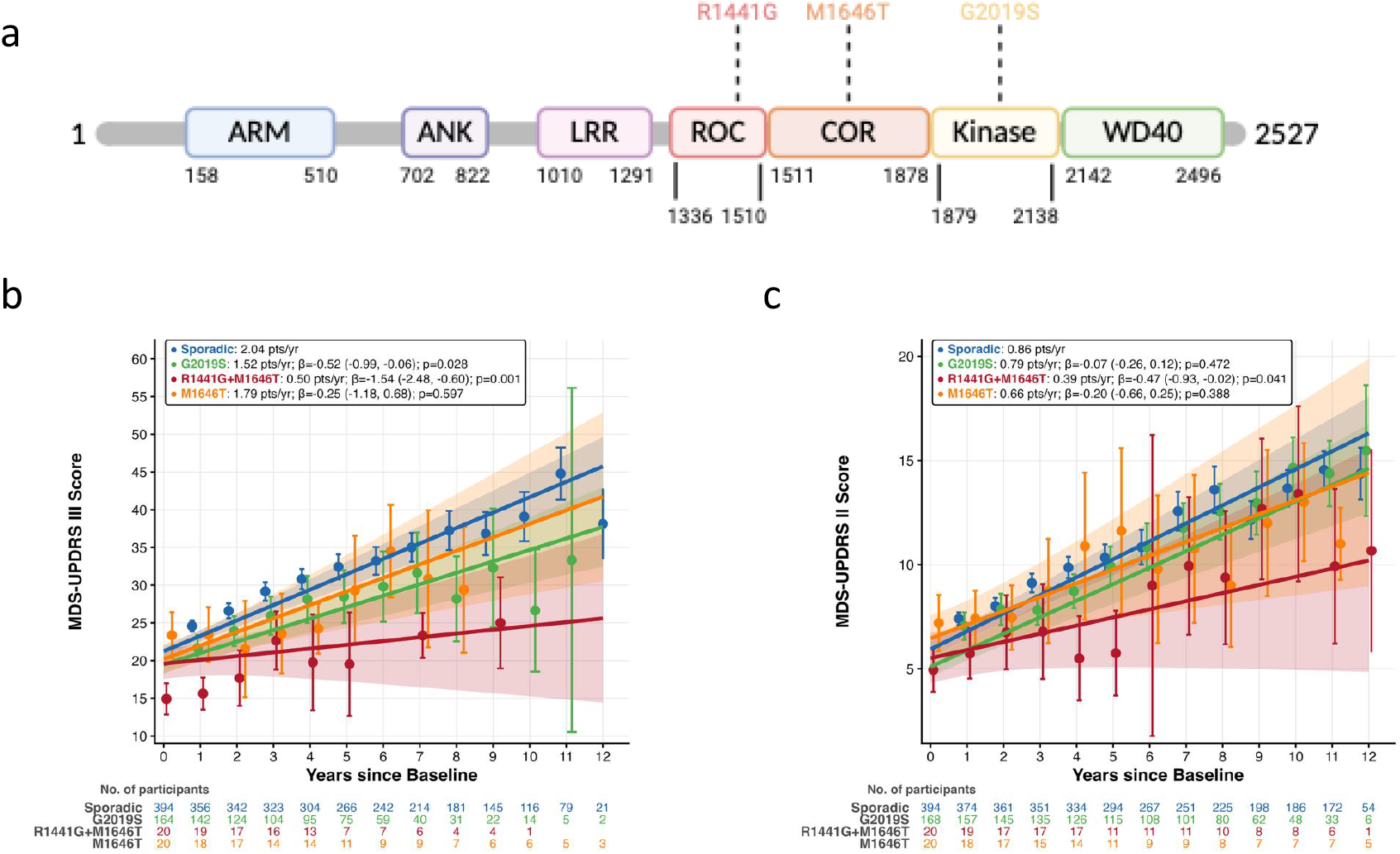
LRRK2 structure and motor symptom progression. **a)** Structure of the *LRRK2* gene. **b)** Change in MDS-UPDRS III unmedicated/OFF-state adjusted for age at onset, disease duration at baseline, sex, race, baseline MDS-UPDRS III score, and levodopa equivalent daily dose. c) Change in MDS-UPDRS II adjusted for age at onset, sex, race, and baseline MDS-UPDRS II score. Lines represent the predicted value over time after adjustment. Data points represent the unadjusted mean observed value at the corresponding time point. Shading and error bars represent 95% confidence intervals for observed and predicted values. All statistics are derived from the adjusted model. Abbreviations: PD, Parkinson’s disease; *LRRK2*, Leucine-rich repeat kinase 2; MDS-UPDRS III, Movement Disorder Society Unified Parkinson’s Disease Rating Scale Part III; ARM, Armadillo repeat; ANK, Ankyrin repeat; LRR, Leucine-rich repeat; ROC, Ras of complex protein; COR, C-terminal of ROC; WD40, tryptophan–aspartic acid repeat.

## Methods and Results

We analyzed up to 12 years (mean follow-up=6.3 years) of Movement Disorder Society Unified Parkinson’s Disease Rating Scale (MDS-UPDRS) Parts I, II (secondary outcome), III (primary outcome), and IV, Montreal Cognitive Assessment (MoCA), as well as up to 5 years (mean follow-up=3.8 years) of dopamine transporter imaging (DAT-SPECT) from 603 PPMI participants with PD and available whole-genome sequencing data, including 394 with sporadic PD, 169 G2019S carriers, 20 R1441G+M1646T carriers, and 20 M1646T carriers.^6^ Participants with *GBA, SNCA, PRKN, PINK1, PARK7*, and *VPS35* mutations were excluded. Demographics and baseline clinical characteristics for each group are located in Supplemental Table 1.

Associations between genotype and outcome measures were assessed using linear mixed-effects models including genotype, time since baseline, and their interaction, adjusted for age at onset, disease duration at baseline, sex, race, and the corresponding baseline outcome value, with continuous covariates centered at their means; motor outcomes were additionally adjusted for concurrent levodopa equivalent daily dosage (LEDD), centered at zero. Models included participant-level random intercepts and slopes for time from baseline visit with an unstructured covariance matrix and allowed for group-specific residual variance. Each outcome measure was modeled independently. Pairwise contrasts between carrier groups were evaluated with Wald tests. Analyses were performed in R version 4.4.1 (nlme package). A significance level of α=0.05 was used for all analyses.

R1441G+M1646T (76% slower; 0.50 vs. 2.04 points/year; β=−1.54 [95% CI: −2.48, −0.60]; p=0.001) and G2019S carriers (26% slower; 1.52 points/year; β=−0.52 [−0.99, −0.06]; p=0.03) exhibited slower progression in clinician-reported motor symptoms (unmedicated/OFF-state MDS-UPDRS III) than participants with sporadic PD, while M1646T carriers did not (1.79 points/year; β=−0.25 [−1.18, 0.68]; p=0.60; Figure 1b; Supplemental Table 2). Results were robust after adjusting for α-synuclein seed amplification assay (SAA) status and after restricting follow-up to 5 years (Supplemental Table 3). R1441G+M1646T carriers progressed more slowly than G2019S carriers (β=−1.02 [−2.01, −0.03]; p=0.04) and M1646T carriers (β=−1.29 [−2.59, 0.00]; p=0.05; Supplemental Table 3).

Subscore analyses revealed a lack of orofacial symptom progression (−0.09 vs. 0.12 points/year; β=−0.21 [−0.28, −0.13]; p<0.001) in R1441G+M1646T carriers, with attenuated bradykinesia (64% slower; β=−0.68 [−1.17, −0.18]; p=0.008) and axial function decline (76% slower; β=−0.29 [−0.48, −0.11]; p=0.002). G2019S carriers also exhibited attenuated bradykinesia (26% slower; β=−0.28 [−0.52, −0.03]; p=0.03) and axial function decline (26% slower; β=−0.10 [−0.19, −0.01]; p=0.03; Supplemental Table 2).

R1441G+M1646T carriers also exhibited 55% slower progression in self-reported motor symptoms (MDS-UPDRS II; 0.39 vs. 0.86 points/year; β=−0.47 [−0.93, −0.02]; p=0.04) while G2019S (0.79 points/year; β=−0.07 [−0.26, 0.12]; p=0.47) and M1646T carriers (0.66 points/year; β=−0.20 [−0.66, 0.26]; p=0.39) did not (Figure 1c; Supplemental Table 2). Motor complications (MDS-UPDRS IV), non-motor decline (MDS-UPDRS I), cognitive decline (MoCA), and dopaminergic imaging (DAT-SPECT) were directionally consistent but varied in statistical significance (Supplemental Table 4).

## Discussion

Our findings expand upon previously reported heterogeneity in *LRRK2*-associated PD.^1–3^ R1441G+M1646T and G2019S carriers progressed more slowly than sporadic PD while M1646T carriers did not. While less common than G2019S, R1441G+M1646T carriers showed the largest reduction in motor symptom progression relative to sporadic PD.

R1441G has been traced to a seventh-century founder event in Northern Spain and co-segregates with M1646T on a shared ∼5.8 Mb haplotype.^4,5^ Unlike G2019S, which is located in the kinase domain and increases *LRRK2* kinase activity directly,^7^ R1441G is located in the Ras-of-complex (ROC) domain where it impairs GTPase hydrolysis and indirectly increases kinase activity.^7^ M1646T, in the adjacent C-terminal of ROC (COR) domain, is also associated with increased kinase activity and PD risk.^4,5,8^ Whether increased kinase activity underlies the slower progression observed in R1441G+M1646T carriers requires further investigation.

A previous cross-sectional study found no difference in MDS-UPDRS or MoCA between R1441G+M1646T and G2019S carriers in the PPMI cohort. Here we found that R1441G+M1646T carriers progressed more slowly than G2019S carriers, a difference cross-sectional comparisons are not designed to capture.^3^ Our finding of slower progression in G2019S carriers aligns with previous longitudinal analyses of the PPMI cohort.^9^

We previously reported that a trend toward slower progression in SAA-negative LRRK2 PD was driven by R1441G+M1646T carriers.^10^ Here, slower progression persisted after adjusting for SAA status. However, genotype and SAA positivity are strongly associated, ranging from 91% in sporadic PD and 95% in M1646T carriers to 55% in G2019S carriers and 25% in R1441G+M1646T carriers, and statistical adjustment alone cannot fully disentangle their contributions.

The R1441G+M1646T and M1646T groups were small (n=20 each), limiting statistical power for comparisons among carrier groups. R1441G+M1646T carriers progressed more slowly than M1646T carriers; however, this comparison only reached borderline significance. R1441G has not been observed independently of M1646T in any sequenced cohort to date, and available data are predominantly of European ancestry; whether the two variants dissociate in other populations remains unknown.^5^ Distinguishing the individual contributions of R1441G and M1646T will require larger cohorts and functional studies.

## Data Availability

Data used in the preparation of this article are openly available to qualified researchers upon registering for access via the PPMI website. Data were obtained in July 2026 from the PPMI database (www.ppmi-info.org/access-data-specimens/download-data, RRID:SCR_006431). A list of all data used, and the code generated in this study can be found at https://github.com/jacksongsch/lrrk2_progression.

## Abbreviations

COR: C-terminal of ROC
LRRK2: Leucine-rich repeat kinase 2
PD: Parkinson’s disease
PPMI: Parkinson’s Progression Markers Initiative
ROC: Ras of complex protein
SAA: Seed amplification assay

## Declarations

### Ethics approval and consent to participate

This study uses deidentified demographic and clinical data from PPMI. Human subjects research is exempt as defined by Title 45 Code of Regulations (CFR)46.

### Consent for publication

Not applicable.

### Competing interests

The authors declare no conflicts of interest. Funding

This research was funded in part by Aligning Science Across Parkinson’s grants ASAP-000312 and MJFF-028544 through the Michael J. Fox Foundation for Parkinson’s Research (MJFF) and by the National Institutes of Health through the National Institute of Neurological Disorders and Stroke grant R01NS102735.

### Authors’ contributions

Conceptualization: JGS and XC; Data Curation: JGS; Methodology, Software, and Formal Analysis: JGS, XZ, JW, and XC; Visualization: JGS; Data Validation: JGS, XZ, JW, and XC; Writing – Original Draft: JGS and XC; Writing – Review & Editing: JGS, XZ, JW, and XC; Supervision: XC; Funding Acquisition: XC. JS and XC have direct access to and have verified the underlying data. All authors have read and approved the final version of the manuscript.

## Acknowledgements

The authors would like to thank PPMI – a public-private partnership – funded by the Michael J. Fox Foundation for Parkinson’s Research and funding partners, including 4D Pharma, Abbvie, AcureX, Allergan, Amathus Therapeutics, Aligning Science Across Parkinson’s, AskBio, Avid Radiopharmaceuticals, BIAL, BioArctic, Biogen, Biohaven, BioLegend, BlueRock Therapeutics, Bristol-Myers Squibb, Calico Labs, Capsida Biotherapeutics, Celgene, Cerevel Therapeutics, Coave Therapeutics, DaCapo Brainscience, Denali, Edmond J. Safra Foundation, Eli Lilly, Gain Therapeutics, GE HealthCare, Genentech, GSK, Golub Capital, Handl Therapeutics, Insitro, Jazz Pharmaceuticals, Johnson & Johnson Innovative Medicine, Lundbeck, Merck, Meso Scale Discovery, Mission Therapeutics, Neurocrine Biosciences, Neuron23, Neuropore, Pfizer, Piramal, Prevail Therapeutics, Roche, Sanofi, Servier, Sun Pharma Advanced Research Company, Takeda, Teva, UCB, Vanqua Bio, Verily, Voyager Therapeutics, the Weston Family Foundation and Yumanity Therapeutics.

## Notes

### Competing Interest Statement

The authors have declared no competing interest.

## References

1 Healy DG, Falchi M, O’Sullivan SS, et al. Phenotype, genotype, and worldwide genetic penetrance of LRRK2-associated Parkinson’s disease: a case-control study. Lancet Neurol. 2008 Jul;7(7):583–90. doi: 10.1016/S1474-4422(08)70117-0. Epub 2008 Jun 6. PMID: 18539534; PMCID: PMC2832754.

2 Taymans JM, Fell M, Greenamyre T, et al. Perspective on the current state of the LRRK2 field. NPJ Parkinsons Dis. 2023 Jul 1;9(1):104. doi: 10.1038/s41531-023-00544-7 PubMed PMID 37393318; PMCID: PMC10314919.

3 Hadad R, Alcalay RN, Senderova I, et al. Parkinson’s disease associated with LRRK2-R1441C mutation: Characterization and comparison with other LRRK2 mutations. J Parkinsons Dis. 2025 Aug;15(5):1029–1034. doi: 10.1177/1877718X251354986. Epub 2025 Jul 4. PMID: 40611668; PMCID: PMC13347503.

4 Mata IF, Hutter CM, González-Fernández MC, et al. Lrrk2 R1441G-related Parkinson’s disease: evidence of a common founding event in the seventh century in Northern Spain. Neurogenetics. 2009 Oct;10(4):347–53. doi: 10.1007/s10048-009-0187-z. Epub 2009 Mar 24. PMID: 19308469; PMCID: PMC2821036.

5 Bryant N, Malpeli N, Ziaee J, et al. Identification of LRRK2 missense variants in the accelerating medicines partnership Parkinson’s disease cohort. Hum Mol Genet. 2021 Apr 30;30(6):454–466. doi: 10.1093/hmg/ddab058 PubMed PMID 33640967; PMCID: PMC8101351.

6 Marek K, Chowdhury S, Siderowf A, et al. The Parkinson’s progression markers initiative (PPMI) - establishing a PD biomarker cohort. Ann Clin Transl Neurol. 2018 Oct 31;5(12):1460–1477. doi: 10.1002/acn3.644 PubMed PMID 30564614; PMCID: PMC6292383.

7 Goveas L, Mutez E, Chartier-Harlin MC, Taymans JM. Mind the Gap: LRRK2 Phenotypes in the Clinic vs. in Patient Cells. Cells. 2021 Apr 22;10(5):981. doi: 10.3390/cells10050981 PubMed PMID 33922322; PMCID: PMC8145309.

8 Sosero YL, Yu E, Krohn L, et al. LRRK2 p.M1646T is associated with glucocerebrosidase activity and with Parkinson’s disease. Neurobiol Aging. 2021 Jul;103:142.e1-142.e5. doi: 10.1016/j.neurobiolaging.2021.02.018. Epub 2021 Feb 28 PubMed PMID 33781610; PMCID: PMC8178224.

9 Sun X, Dou K, Xue L, et al. Comprehensive analysis of clinical and biological features in Parkinson’s disease associated with the LRRK2 G2019S mutation: Data from the PPMI study. Clin Transl Sci. 2024 Jan;17(1):e13720. doi: 10.1111/cts.13720. PMID: 38266062; PMCID: PMC10804919.

10 Schumacher JG, Zhang X, Macklin EA, et al. Baseline α-synuclein seeding activity and disease progression in sporadic and genetic Parkinson’s disease in the PPMI cohort. EBioMedicine. 2025 Sep;119:105866. doi: 10.1016/j.ebiom.2025.105866. Epub 2025 Aug 6. PMID: 40773926; PMCID: PMC12354789.

